# Prospective real-world evidence of mental health outcomes following supervised psilocybin services within Oregon’s state-regulated model

**DOI:** 10.64898/2026.02.18.26346580

**Authors:** Amanda Gow, Emily Shih, Ryan Reid, Jimmy J. Qian, Charan Mellor, Lynne Alison McInnes, Robin Carhart-Harris, Jennie N. Davis

**Affiliations:** Bendable Therapy PO Box 1229, Bend, OR 97709, USA; Osmind 3130 20th Street, Suite 250, San Francisco, CA 94110, USA; Weill Institute for Neurosciences, University of California San Francisco 1651 4th St, San Francisco, CA 94158, USA; Center for Health Systems Effectiveness, Oregon Health & Science University 3030 S Moody Ave Unit 200, Portland, OR 97239, USA

**Keywords:** psilocybin, Oregon, mental health, depression, anxiety, well-being, psychedelics

## Abstract

**Background:** In 2020, Oregon established the first regulated framework for adult psilocybin services using naturally-derived mushroom products. No studies have examined mental health outcomes among individuals receiving psilocybin in this real-world, state-regulated context.

**Aims:** To evaluate the effects of supervised psilocybin services under Oregon’s model on self-reported symptoms of depression, anxiety, and well-being, and document session-related aftereffects and doses consumed.

**Methods:** This was a prospective, naturalistic study (March 2024-April 2025) among adults ≥21 years participating in a legal psilocybin services program. Online surveys were completed pre-session, 1-day, and 30-days post-session. Primary outcomes were change in depression, anxiety, and well-being symptoms pre-session to 30-days post-session evaluated using linear mixed-effects models (random effect: participant; fixed effects: timepoint, sex, concurrent psychiatric medication use, age, dose [total psilocybin equivalents, TPE]). Post-session visual perceptual or psychological aftereffects were assessed at follow-up.

**Results:** Participants (n=88; median age 43 years; 52% male) were predominantly Oregon residents (53.4%), psychedelic-experienced (64.8%), and using psychiatric medication (46.6%). All outcomes showed significant within-person improvements 30-days post-session (p<0.001), including in sensitivity analyses stratified by concurrent psychiatric medication usage (p<0.001 all outcomes, both groups). Two participants (2.3%) reported session-related distressing visual perceptual aftereffects 1-day post-session, resolved at 30-days; three (3.4%) reported psychological aftereffects at 30-days. Mean dose was 27.8 mg (SD=8.2) TPE.

**Conclusions:** Psilocybin sessions delivered under Oregon’s regulatory model were associated with significant improvements in depression, anxiety, and well-being. As other U.S. jurisdictions explore similar models, this initial real-world evidence supports the potential utility of legal psilocybin services for improved mental health.

## Introduction

Psilocybin has emerged as a promising therapeutic intervention for mental health disorders, including depression and anxiety (Carhart-Harris et al., 2016, 2018; Dorczok et al., 2025; Goodwin et al., 2022; Goodwin, Aaronson, et al., 2023; Griffiths et al., 2016; Khan et al., 2026). As a naturally occurring psychoactive alkaloid found in certain species of mushrooms, once ingested, psilocybin is converted to psilocin to produce its characteristic psychedelic effects by acting on serotonin receptors (e.g., 5-HT2_A_) (Passie et al., 2002). These neuropharmacological actions are thought to underlie the improvements in mental health symptoms observed across controlled clinical trials and naturalistic observational studies (Calnan et al., 2025; Carhart-Harris et al., 2021; Compass, 2025; Davis et al., 2021; de la Salle et al., 2024; Goodwin et al., 2022; Goodwin, Aaronson, et al., 2023; Griffiths et al., 2016; Irrmischer et al., 2025; Nygart et al., 2022; Raison et al., 2023; Ross et al., 2016; von Rotz et al., 2023).

Most psilocybin clinical trials enroll highly selective samples under tightly controlled conditions, typically excluding individuals with complex psychiatric diagnoses or active psychiatric medication use, and administering synthetic psilocybin within standardized dosing protocols (Carhart-Harris et al., 2021; Davis et al., 2021; Griffiths et al., 2016; Raison et al., 2023; Ross et al., 2016; von Rotz et al., 2023). Only one trial has used a natural mushroom formulation to examine impacts of psilocybin on alcohol use disorder, with findings highlighting the therapeutic potential of natural mushroom formulations (Jensen et al., 2025). Naturalistic studies introduce greater variability in participants’ medical and psychiatric histories, session formats, and psilocybin source (often whole fruit mushrooms), yet still demonstrate improvements in depression, anxiety, and well-being (Calnan et al., 2025; de la Salle et al., 2024; Irrmischer et al., 2025; Morton et al., 2023; Nygart et al., 2022; Tap et al., 2025). However, additional research that bridges controlled clinical efficacy trials to real-world settings is needed to understand psilocybin’s mental health benefits in diverse, community-based populations.

In 2020, Oregon voters approved Measure 109, establishing the first regulated psilocybin services model in the United States and globally. These services are administered by the Oregon Health Authority (OHA) through its Oregon Psilocybin Services (OPS) section (Oregon Health Authority, n.d.) Oregon’s non-medical model permits adults aged 21 years and older to access psilocybin by consuming products derived from whole fruit mushrooms (*Psilocybe cubensis*) at licensed service centers under the supervision of licensed facilitators (Oregon Health Authority, 2024). The model does not require a medical diagnosis, prescription, or clinical referral, despite mental health applications being common reasons for psilocybin use (Dorczok et al., 2025). In 2025, statewide aggregate data released under Senate Bill 303 began providing basic information, including demographics, psilocybin dosages, and session types. However, these data do not include clinical outcomes or follow-up assessments, limiting insight into the model’s therapeutic effectiveness.

To address this gap, this study aimed to evaluate the effects of supervised psilocybin services under Oregon’s model on self-reported symptoms of depression, anxiety, and well-being, and document session-related aftereffects and psilocybin doses consumed. Predictors of session dosage were additionally explored to understand potential correlates of dosing decisions (e.g., participant characteristics, mental health history). Characterizing who uses psilocybin services and with what outcomes is critical to understanding the benefits and limitations of Oregon’s regulated framework.

## Methods

### Setting and Participants

This prospective, naturalistic study (March 2024-April 2025) was integrated into an existing Oregon legal psilocybin services program (“study site”) (**Figure 1**). Oregon eligibility requirements for legal psilocybin services are: aged 21 years or older, no lithium use in the past 30 days, no active psychosis, and no active suicidal or homicidal ideation (Oregon Health Authority, 2024). Oregon regulatory minimums require one facilitator-client preparation session 24-hours in advance of a psilocybin administration session (where eligibility criteria are assessed) and to offer an optional integration session post-administration (Oregon Health Authority, n.d.). In addition to the Oregon eligibility criteria, the study site enhanced the Oregon requirements by including additional eligibility questions in their screening application (not pregnant or breastfeeding, no uncontrolled hypertension or other clinically significant cardiovascular conditions requiring further medical evaluation (Nahlawi et al., 2025)); requiring a virtual interview where a study site staff member reviewed all Oregon and study site eligibility criteria with the individual; and offering multiple post-session support sessions (Figure 1). During the study period, all individuals who attended the virtual interview and met both Oregon and study site eligibility criteria were invited to the study and to complete the written informed consent. Study participants received no compensation for participation. The study site independently offers scholarships to some clients to offset the cost of psilocybin services; some participants received these scholarships, but scholarship status did not influence study enrollment. WCG Institutional Review Board approved this study as human subjects research.

**Figure 1.**
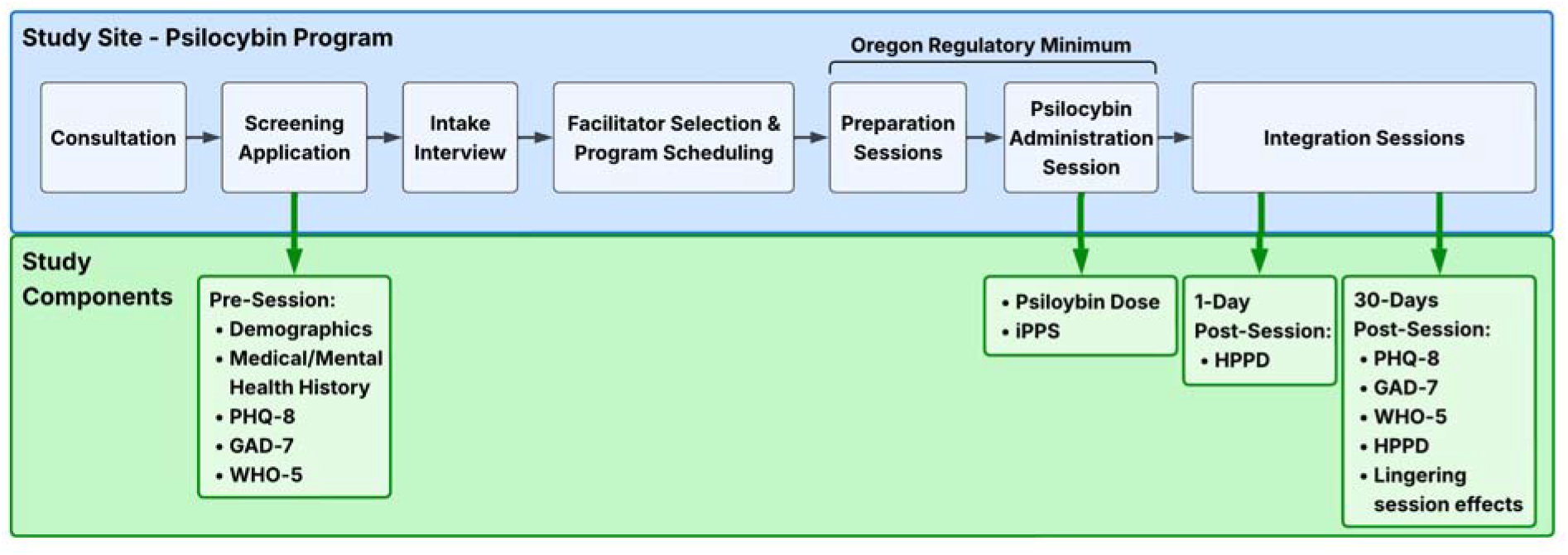
Study components within the framework of a legal psilocybin services program in Oregon. Study components were integrated into an existing legal psilocybin services program in Oregon (study site). Oregon regulatory minimums require one facilitator-client preparation session 24-hours in advance of a psilocybin administration session and to offer an optional integration session post-administration. The study site psilocybin program included additional comprehensive screening, preparation sessions, and structured integration support sessions. Note that mental health outcomes (PHQ-8, GAD-7, WHO-5) were also assessed at the 1-day follow-up but are reported only as a supplementary analysis because the instruments’ 2-week recall periods included both before and after the administration session; results are included in Supplemental Table 2. Abbreviations: GAD-7, Generalized Anxiety Disorder-7 questionnaire; HPPD, Hallucinogen Persisting Perception Disorder; PHQ-8, Patient Health Questionnaire-8; iPPS, Imperial Psychedelic Predictor Scale; WHO-5, World Health Organization Well-Being-5 Index.

### Measures and procedures

#### Pre-psilocybin session measures and procedures

A pre-session screening application collected demographics, medical, mental health and psychedelic use histories, medication usage, and both state and study site eligibility screening questions (e.g., lithium usage, psychosis, suicidality, pregnancy/breastfeeding). Validated scales assessed self-reported symptoms of depression (Patient Health Questionnaire, PHQ-9), anxiety (Generalized Anxiety Disorder, GAD-7), and well-being (World Health Organization Well-Being Index, WHO-5) (Figure 1).

The PHQ-9 maps nine depression symptoms over the past two weeks onto the Diagnostic and Statistical Manual of Mental Disorders, Fourth Edition, Text Revision criteria for major depressive disorder; items are scored on a 4-point Likert scale (0=“not at all” to 3=“nearly every day”) (Kroenke et al., 2001)). For this study, the PHQ-9 was modified to PHQ-8 by excluding the suicidal ideation item. As suicidal ideation is criterion for exclusion from psilocybin services (Oregon Health Authority, 2024), and suicidality was assessed in a separate dedicated screening question in the screening application, the PHQ-9 suicidal ideation item was considered redundant for this population and was removed. PHQ-8 scores were adjusted according to established procedures (Shin et al., 2019). The GAD-7 is a 7-item self-report scale assessing generalized anxiety severity over the past two weeks with items rated on a 4-point Likert scale (0=“not at all” to 3=“nearly every day”) (Spitzer et al., 2006). The WHO-5 is a 5-item measure of subjective psychological well-being where participants rated positive mood states (e.g., feeling cheerful, relaxed, or energetic) over the past two weeks on a 5-point scale (0=“at no time” to 5=“all of the time”) (Topp et al., 2015).

#### Day-of psilocybin session measures and procedures

On the administration session day, before consuming psilocybin, participants completed the Imperial Psychedelic Predictor Scale (iPPS), a validated 8-(individual sessions) or 9-item (group sessions) multidimensional self-report instrument designed to predict acute features of the psychedelic experience across the domains of set, intention, and rapport; items were rated 0-100 (Angyus et al., 2024). Consumed psilocybin and psilocin doses (mg) were recorded.

#### Post-psilocybin session measures and procedures

Post-session surveys were emailed to participants 1-day and 30-days post session. Both surveys included follow-up PHQ-8 (post-session surveys also excluded the suicidal ideation item due to concerns that these surveys were reviewed by researchers, not clinicians, and on timelines that may not have been prompt enough to respond to reports of self-harm (Shin et al., 2019)), GAD-7, and WHO-5 assessments. The surveys also assessed visual perceptual aftereffects potentially associated with Hallucinogen Persisting Perception Disorder (HPPD), a condition of enduring, distressing visual perceptions post-psychoactive substance intake (Figure 1). HPPD visual perceptual aftereffects attributed to the psilocybin session and not due to other medical or mental health conditions may signify risk for an HPPD (Zhou et al., 2025). The 30-day post-session survey also collected data on integration session attendance (a post-session participant-facilitator meeting to discuss and process the psilocybin session), and lingering unpleasant psychological effects attributed to the psilocybin session (i.e., anxiety, existential struggle, social disconnection, depersonalization/derealization) (Schlag et al., 2022). The occurrence of acute adverse behavioral and medical reactions or emergency service calls during the psilocybin session were required reporting by service centers to OHA under ORS 475A.372 (Oregon Legislature, 2025) and OAR 333-333-4700 (Oregon Health Authority, 2025), and related publicly-available data are reported. For participants completing two sessions within the follow-up period (e.g., weeklong retreats), data were collected according to the first session only. Online assessments were completed via Jotform (Jotform Inc.). **Appendix A (Supplemental Table 1)** contains a summary of the study’s validated scales.

#### Psilocybin session procedures and dosing

Participants completed individual or group facilitator-supervised psilocybin sessions at licensed service centers following Oregon regulations (Oregon Health Authority, n.d.), and the study site’s psilocybin program protocols were not altered for this study. Psilocybin was consumed as whole mushrooms or tea made with ground whole mushrooms. Session dosing followed the study site’s protocol of a total dose of 20-30 mg psilocybin including optional ‘booster doses,’ or additional offered approximately 1-hour after the initial dose. Per Oregon rules, participants may request lower or higher doses (up to 50 mg) (Oregon Psilocybin Services, 2025, n.d.).

### Statistical analysis

Participant pre-session characteristics are presented as medians (range) or frequencies (proportions). Primary outcomes were change in mental health status from pre-session to 30-days post-session using the PHQ-8, GAD-7, and WHO-5 scales. Outcomes from the 1-day post-session assessment are not included as primary outcomes because these instruments have a two-week recall period which spanned both before and after the administration session and may reflect a mixture of participant pre- and post-session symptoms. One-day post-session results are therefore presented as a supplementary analysis.

Scores for the PHQ-8 (0-24) were summed, and cut-points of 5, 10, 15, and 20 denoted mild, moderate, moderately severe, and severe depression respectively; scores ≥10 indicated clinically significant depression (Kroenke et al., 2001). GAD-7 scores (0-21) were summed with cut-points of 5, 10, and 15 denoting mild, moderate, and severe anxiety respectively, with ≥10 indicating clinically significant anxiety (Spitzer et al., 2006). WHO-5 scores (0–25) were summed with scores <13 indicating poor well-being. The PHQ-8, GAD-7, and WHO-5 demonstrated strong internal consistency measured with Cronbach’s alpha (PHQ-8: =0.87 pre-session and 30-days post-session; GAD-7: α=0.91 pre-session and α=0.89 30-days post-session; WHO-5: α=0.90 pre-session and α=0.93 30-days post-session).

Per Oregon regulations, mushrooms were grown, processed, and potency-tested for analyte content (psilocybin and psilocin) in Oregon licensed facilities; product packaging listed analyte content in milligrams per gram of whole mushroom (Oregon Psilocybin Services, n.d.). Oregon regulations define dose by psilocybin content; because psilocybin rapidly converts to its pharmacologically active form of psilocin once ingested (Passie et al., 2002), the study site calculates dose as Total Psilocybin Equivalents (TPE = psilocybin mg + 1.39 × psilocin mg), a metric assuming complete conversion of psilocybin to psilocin based on molecular weight (MacCallum et al., 2022). We report both TPE and psilocybin dose.

We used separate linear mixed effects models using the ‘lme4’ package in R to evaluate changes in outcomes across two timepoints (means, SE). Timepoint was modeled as a categorical variable (pre-session as reference), with fixed effects of birth sex (male/female), age (years), concurrent psychiatric medication use (yes/no), and dose (TPE, mg). Participant-level random intercepts accounted for individual differences in pre-session symptom levels and change over time. Prior to analyses, we evaluated covariate collinearity and excluded the session type variable (group vs. individual) due to strong correlation with TPE. Post-session effect sizes (relative to pre-session) were calculated using Cohen’s *d*. This was calculated by dividing the estimated marginal mean differences by the pooled standard deviation, which incorporated both the residual variance and the random intercept variance from the fitted model. Effect sizes were classified as small (0.20-0.49), moderate (0.50-0.79), or large (≥0.80) (Cohen, 1992). Results are reported as estimated marginal means with standard errors, mean differences with 95% confidence intervals, and Cohen’s d. A simulation-based power analysis (□=0.05) indicated approximately 100% power for large effects and 89% for small effects.

### Sensitivity and exploratory analyses

In a sensitivity analysis, we repeated the adjusted models substituting TPE with psilocybin dosage to assess robustness to an alternative dose specification. In a second analysis to assess whether findings were driven by participants with additional dosing, we restricted the sample to participants who completed a single session during the study period (n=75), excluding those with multiple sessions (n=13). In a third analysis, we stratified the sample by concurrent psychiatric medication use and repeated the adjusted models separately among participants with (n=41) and without (n=47) pharmacotherapy to descriptively assess whether patterns of outcomes were consistent across medication status.

To explore correlates of dosing decisions, we used univariate linear regression models to examine predictors of session dose (TPE). Predictors included pre-session PHQ-8, GAD-7, and WHO-5 scores; age; birth sex; most commonly reported diagnoses: depression, anxiety, post-traumatic stress disorder (PTSD); concurrent psychiatric medication use; prior psychedelic experience (Irrmischer et al., 2025). In a separate multivariate regression model, iPPS subscales were entered simultaneously to assess their independent contributions to TPE. Given these analyses’ exploratory nature, p-values were not adjusted for multiple comparisons.

Analyses were conducted using R/R Studio version 4.4.2 (R Foundation), and included only participants who completed all pre- and post-session surveys; we set statistical significance at p<0.05.

### Results

#### Participant characteristics

Descriptive participant characteristics can be found in **Table 1**. During the study period, n=311 screening applications were received. Of these, n=126 individuals met eligibility criteria for psilocybin services and were invited to participate (n=35 did not proceed with psilocybin services during the study period due to self-selected factors, such as financial constraints, scheduling, or other personal reasons), n=91 enrolled (n=3 lost to follow-up), and n=88 completed all study components and comprised the analytic sample. Participants (n=88) ranged in age from 22-79 years (median: 43 years), were approximately balanced by birth sex and gender identity (n=46, 52.0% male; n=44, 50.0% male-identifying), and were predominantly White (n=77, 87.5%), educated (n=74, 84.1% with higher education), and employed (n=59, 67.1%), with only 22.7% (n=20) reporting annual household income below $50,000. Most participants were Oregon residents (n=47, 53.4%) and 15.9% (n=14) were military veterans. Depression (n=45, 51.1%), anxiety (n=37, 42.0%), and PTSD (n=17, 19.3%) were the most frequently reported mental health diagnoses. Almost half (n=41, 46.6%) reported concurrent use of psychiatric medications (medication type and dosage were not systematically collected) and participation in mental health therapy (n=50, 56.8%). The majority (n=57, 64.8%) had prior psychedelic experience, and cost was the most frequently reported barrier to seeking psilocybin services (n=41, 46.6%) (Table 1). Most participants completed individual sessions (n=50, 56.8%) rather than group sessions (n=38, 43.2%), and 80.0% (n=70) completed at least one integration session during follow-up.

**Table 1:** Participant demographic, mental health, and psychedelic experience characteristics.

| Demographic Characteristics | Session Participants (n=88) |
| --- | --- |
| Age, years (median, range) | 43 (22, 79) |
| Birth sex, No. (%) |  |
| Male | 46 (52.0) |
| Female | 41 (46.6) |
| Gender, No. (%) |  |
| Male | 44 (50.0) |
| Female | 40 (45.5) |
| LGBTQIA+ | 3 (3.4) |
| Race<sup>a</sup>, No. (%) |  |
| White | 77 (87.5) |
| American Indian or Alaska Native | 1 (1.1) |
| Mixed/multiple races | 3 (3.4) |
| Ethnicity, No. (%) |  |
| Non-Hispanic/Non-Latinx | 77 (87.5) |
| Hispanic-Latinx | 5 (5.7) |
| Level of education completed, No. (%) |  |
| Graduate or professional degree | 31 (35.2) |
| College or baccalaureate degree | 32 (36.4) |
| Associate's or technical degree | 11 (12.5) |
| High school or some college | 13 (14.8) |
| Employment, No. (%) |  |
| Employed (full-time, part-time, self-employed, or sporadically) | 59 (67.1) |
| Retired | 17 (19.3) |
| Unemployed/on disability | 9 (10.2) |
| Student | 1 (1.1) |
| Annual household income, No. (%) |  |
| ≥ \$100,000 | 30 (34.1) |
| \$50,000-\$99,999 | 25 (28.4) |
| \$10,000-\$49,000 | 18 (20.4) |
| < \$10,000 | 2 (2.3) |
| Decline to Answer | 13 (14.8) |
| Scholarship recipient<sup>b</sup> | 51 (58.0) |
| Residence, No. (%) |  |
| Living in Oregon | 47 (53.4) |
| Living in other U.S. state | 40 (45.5) |
| Living internationally | 1 (1.1) |
| Military veteran, No. (%) | 14 (15.9) |
| **Mental Health Characteristics** |  |
| Reported mental health diagnosis, No. (%) |  |
| Depression | 45 (51.1) |
| Anxiety | 37 (42.0) |
| PTSD | 17 (19.3) |
| Acute Stress | 6 (6.8) |
| OCD | 4 (4.5) |
| Eating disorder | 2 (2.3) |
| Bipolar disorder<sup>c</sup> | 1 (1.1) |
| Other | 6 (6.8) |
| No mental health diagnosis | 32 (36.4) |
| Currently taking psychiatric medications, No. (%) | 41 (46.6) |
| Currently in therapy, No. (%) | 50 (56.8) |
| **Psychedelic Characteristics** |  |
| Prior psychedelic experience, No. (%) | 57 (64.8) |
| Barriers to seeking psilocybin services, No. (%) |  |
| Cost | 41 (46.6) |
| Availability | 16 (18.2) |
| Time and schedule | 21 (23.9) |
| Limited knowledge and experience of psychedelics | 16 (18.2) |
| Current support systems in place (or not in place) | 6 (6.8) |
| Legal concerns | 4 (4.5) |
| Judgement from family and peers | 4 (4.5) |
| Current mental health concerns | 4 (4.5) |
| No concerns | 28 (31.8) |
| Other | 1 (1.1) |

Notes: Some categories of characteristics may not add to 100% as some participants declined to answer demographic questions and data are missing. All data is from the screening application, which gathered pre-session data (prior to a psilocybin administration session). Abbreviations: LGBTQIA+, Lesbian, Gay, Bisexual, Transgender, Queer, Intersex, Asexual, all other identities not explicitly mentioned; OCD, obsessive compulsive disorder; PTSD, post-traumatic stress disorder.
<sup>a</sup> Race variable was a “select all that apply” field. Participants who selected more than one race were grouped into a “Mixed / Multiple race” option and custom responses that didn’t match the standard categories were grouped into “Other”.
<sup>b</sup> Study participants received no compensation for participation. The study site independently offers scholarships to some clients to offset the cost of psilocybin services; some participants received these scholarships, but scholarship status did not influence study enrollment.
<sup>c</sup> Participants who listed bipolar disorder as a mental health diagnosis in their screening application underwent additional screening. Those who never had a formal diagnosis of bipolar disorder and were otherwise eligible were allowed to continue to a psilocybin session.

#### Mental health outcomes after a psilocybin session

We examined whether participation in a supervised psilocybin session was associated with changes in mental health outcomes. **Table 2** includes the proportion of participants reporting mental health symptoms before and 30-days following the psilocybin session. Pre-session, 69.3% (n=61 of participants reported mild to severe depression symptoms, 67.0% (n=59) reported mild to severe anxiety symptoms, and 53.4% (n=47) reported low or very-low sense of well-being, reflecting clinically significant symptom levels across all measures. At approximately 30-days post-session, improvements were observed across measures, with 61.4% (n=54) of participants reporting minimal depression, 70.5% (n=62) reporting minimal anxiety, and 64.8% (n=57) reporting normal levels of well-being.

**Table 2:** Participant classification by PHQ-8, GAD-7, and WHO-5 categories pre- and post-psilocybin session.

| **Participant Classification Category** | **Pre-Session (n=88)** | **30-days Post-Session (n=88)** |
| --- | --- | --- |
| PHQ-8 (depression), No. (%) |  |  |
| Minimal | 27 (30.7) | 54 (61.4) |
| Mild-Moderate | 45 (51.1) | 31 (35.2) |
| Moderately Severe-Severe | 16 (18.1) | 3 (3.4) |
| GAD-7 (anxiety), No. (%) |  |  |
| Minimal | 29 (33.0) | 62 (70.5) |
| Mild-Moderate | 44 (50) | 25 (28.4) |
| Severe | 15 (17.0) | 1 (1.1) |
| WHO-5 (well-being), No. (%) |  |  |
| Normal | 31 (35.2) | 57 (64.8) |
| Borderline | 10 (11.4) | 9 (10.2) |
| Low-very low | 47 (53.4) | 22 (25.0) |

Notes: Participants completed online surveys that contained the 3 mental health questionnaires (PHQ-8, GAD-7, WHO-5) at two timepoints: pre-session and approximately 30-days post-psilocybin administration session. Pre-session surveys were completed, on average, 75 days prior to the psilocybin administration session; 30-day post-session surveys were completed, on average, 34 days after the psilocybin administration session. Abbreviations: GAD-7, Generalized Anxiety Disorder-7; PHQ-8, Patient Health Questionnaire-8; WHO-5, World Health Organization-5 Well-Being index.

Modeling results similarly reflected significant improvements in mental health outcomes at 30-days post-session, which are depicted in **Figure 2** and **Table 3** (modeling results with outcomes from the 1-day post-session survey are in **Supplemental Table 2**). PHQ-8 and GAD-7 mean scores significantly decreased pre-session to 30-days post-session, suggesting robust declines in depression and anxiety symptoms: PHQ-8 score change of -4.62 (0.57 SE) points; *p*<0.001; *d*=-0.90 (95% CI: -1.12, -0.68) large; GAD-7 score change of -4.85 (0.57) points; *p*<0.001; *d*=-1.04 (-1.28, -0.80) large. WHO-5 well-being scores also improved at follow-up with a score change of +4.26 (0.55) points; *p*<0.001; *d*=0.84 (0.62, 1.05) large), suggesting significant gains in well-being. Of the covariates examined, only concurrent psychiatric medication use was positively associated with post-session PHQ-8 scores (*p*=0.038).

**Figure 2.**
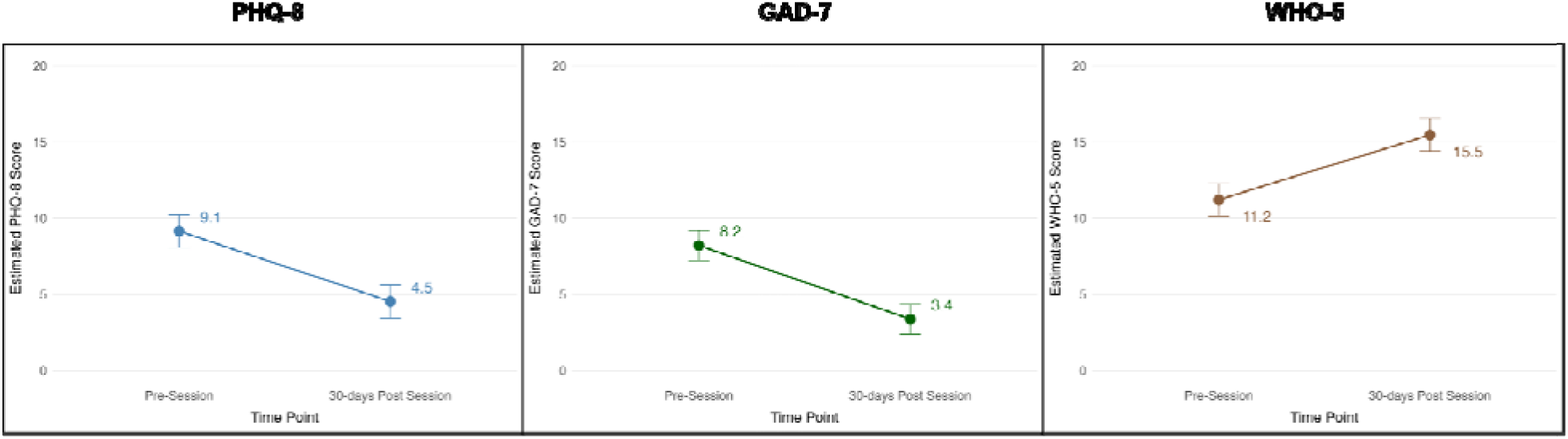
PHQ-8, GAD-7, and WHO-5 estimated marginal mean scores before and 30-days following a legal psilocybin session in Oregon. Change in PHQ-8 (depression), GAD-7 (anxiety), and WHO-5 (well-being) scores are represented as estimated marginal means with error bars indicating standard errors. Adjusted linear mixed effects models evaluated change in outcomes, with all models significant at p<0.001. Abbreviations: GAD-7, Generalized Anxiety Disorder-7; PHQ-8, Patient Health Questionnaire-8; WHO-5, World Health Organization Well-Being Index-5.

**Table 3:** Changes in PHQ-8, GAD-7, and WHO-5 scores from pre-session to 30-days post-psilocybin session (n=88 participants)

| Measure | Pre-session<br>M (SE) | 30-days post-session<br>M (SE) | Change pre-session to post-session<br>M (SE) [95% CI] | p-value | t (df) | Cohen's d<br>[95% CI] |
| --- | --- | --- | --- | --- | --- | --- |
| *Unadjusted* |  |  |  |  |  |  |
| PHQ-8 | 9.15 (0.56) | 4.52 (0.56) | -4.62 (0.57) [-5.77, -3.48] | <0.001 | -8.06 (87) | -0.89 [-1.11, -0.67] |
| GAD-7 | 8.22 (0.51) | 3.36 (0.51) | -4.85 (0.57) [-5.99, -3.71] | <0.001 | -8.48 (87) | -1.02 [-1.26, -0.78] |
| WHO-5 | 11.2 (0.54) | 15.5 (0.54) | +4.26 (0.55) [3.18, 5.35] | <0.001 | 7.80 (87) | 0.83 [0.62, 1.05] |
| *Adjusted* |  |  |  |  |  |  |
| PHQ-8 | 9.15 (0.55) | 4.52 (0.55) | -4.62 (0.57) [-5.77, -3.48] | <0.001 | -8.06 (87) | -0.90 [-1.12, -0.68] |
| GAD-7 | 8.22 (0.50) | 3.36 (0.50) | -4.85 (0.57) [-5.99, -3.71] | <0.001 | -8.48 (87) | -1.04 [-1.28, -0.80] |
| WHO-5 | 11.2 (0.54) | 15.5 (0.54) | +4.26 (0.55) [3.18, 5.35] | <0.001 | 7.80 (87) | 0.84 [0.62, 1.05] |

Notes: Results reported as mean (SE). Covariates included birth sex (male/female/decline to answer), age (years), concurrent psychiatric medication use (yes/no), and total psilocybin equivalents (TPE, mg) as a continuous dosage. Only concurrent psychiatric medication use (pre-session) was positively associated with PHQ-8 scores at 30-days post-session (p=0.038). No covariates were significant in the models with GAD-7 and WHO-5. Cohen's d interpretations: small d=0.20-0.49; moderate d=0.50-0.79; large d≥0.80. Abbreviations: GAD-7, Generalized Anxiety Disorder-7 questionnaire; PHQ-8, Patient Health Questionnaire-8; WHO-5, World Health Organization Well-Being Index.

Sensitivity analyses, one substituting psilocybin for TPE as the dose covariate in all outcomes models (results in **Supplemental Table 3)** and another limited to participants who completed a single administration session (n=75) (results in **Supplemental Table 4**), yielded results consistent with the primary analysis, with no change in the direction or magnitude of timepoint effects. In a third sensitivity analysis, results from stratified models by concurrent psychiatric medication use were largely consistent with those from the full cohort, with significant post-session improvement across all outcomes observed in both groups (p<0.001) **(Supplemental Table 5)**. Overall, these findings support the robustness of the primary results.

The screening application that provided pre-session PHQ-8, GAD-7, and WHO-5 scores was completed a median of 62 days before the administration session (IQR 41.8 to 94.2; range 9 to 291; mean=75.2; SD=51.8). Just under half (46.6%) of participants were assessed within 60 days of the administration session. Cohort assessment distributions were as follows: 11.4% completed pre-session assessment within 30 days of their session, 35.2% between 31 and 60 days, 27.3% between 61 and 90 days, 13.6% between 91 and 120 days, 8.0% between 121 and 180 days, and 4.5% more than 180 days before. The 30-day follow-up survey was completed a median of 33 days after the session (IQR 31 to 38; range 30 to 44; mean=34.8; SD=4.8). A sensitivity analysis examining assessment interval length found no significant associations with pre-session mental health symptoms (all r<0.08; all p>0.48) or with the magnitude of change in outcomes (all r<0.13; all p>0.26).

#### Post-session aftereffects

Participants (n=35, 39.7%) reported visual perception aftereffects at 1-day post-session, with two (2.3%) participants indicating these aftereffects caused significant distress and were attributed to the psilocybin session, putting them potentially at risk for HPPD later on. At 1-day post-session, the most frequently reported visual perceptual aftereffects were “Intensified Colors” (n=19, 21.6%) and “False perceptions of movement in the peripheral visual fields” (n=14, 15.9%) (results in **Supplemental Table 6**). At 30-days post session, the most frequently reported aftereffect was “False perceptions of movement in the peripheral visual fields” (n=7, 8.0%). At 30-days post-session, 16 participants (18.2%) endorsed at least one visual perceptual aftereffect; however, none reported significant distress or impairment. Three (3.4%) participants reported lingering negative psychological aftereffects of the psilocybin experience at 30-days post-session, noting increased feelings of anxiety or fear, existential struggle, social disconnections, and difficulty coping with family members. No adverse behavioral or medical reactions (ORS 475A.372) or emergency services contact reports (OAR 333-333-4700) were reported by service centers for study participants.

#### Session dose and exploratory analyses predicting psilocybin session dose

**Table 4** contains the average session doses measured in TPE and psilocybin. The average dose (TPE) consumed by participants was 27.8 mg (8.2 SD), and was 24.3 mg (8.3 SD) when restricted to psilocybin. In exploratory analyses examining predictors of dose (TPE), the only significant predictor was concurrent use of psychiatric medications, which was associated with greater administered dose (*b*=3.44, *p*<0.05, **Supplemental Table 7)**; these participants received an average dose of 29.6 mg TPE (26.2 mg TPE for those not taking psychiatric medications). No iPPS subscales except preparedness were significantly associated with dose (β=0.20; p=0.04); results can be found in **Supplemental Table 8**.

**Table 4:**
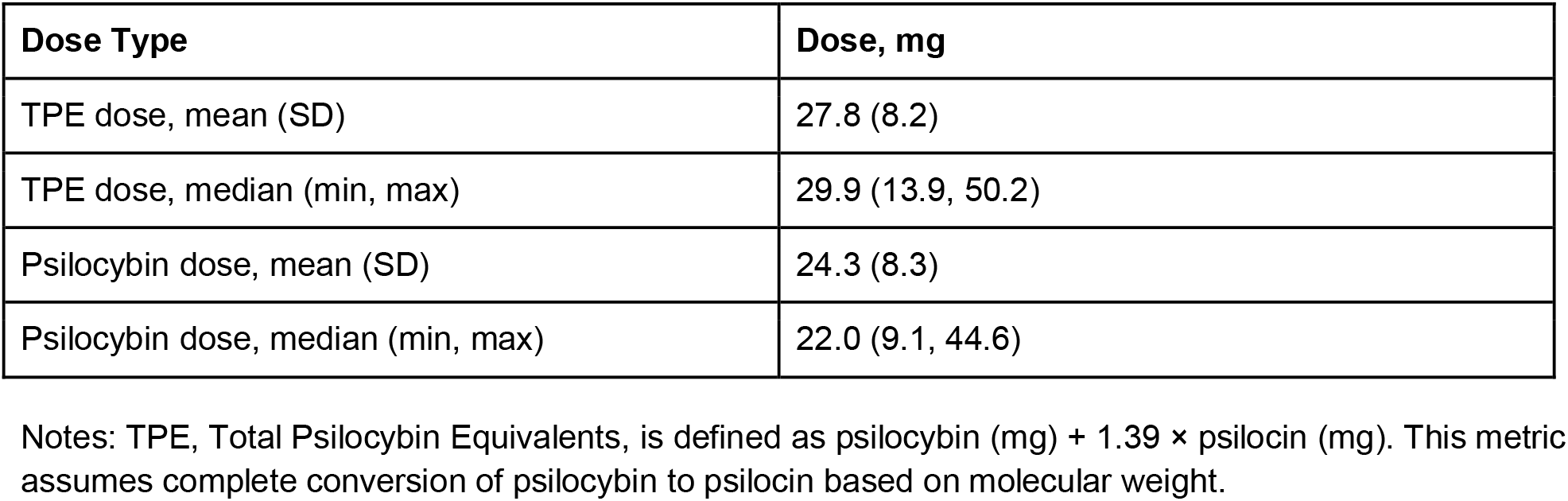
TPE and psilocybin doses consumed by participants during a legal psilocybin session in Oregon.

| Dose Type | Dose, mg |
| --- | --- |
| TPE dose, mean (SD) | 27.8 (8.2) |
| TPE dose, median (min, max) | 29.9 (13.9, 50.2) |
| Psilocybin dose, mean (SD) | 24.3 (8.3) |
| Psilocybin dose, median (min, max) | 22.0 (9.1, 44.6) |

Notes: TPE, Total Psilocybin Equivalents, is defined as psilocybin (mg) + 1.39 × psilocin (mg). This metric assumes complete conversion of psilocybin to psilocin based on molecular weight.

### Discussion

This prospective, naturalistic study presents the first results examining associations between supervised psilocybin services and self-reported mental health symptoms within Oregon’s state-regulated model. We found that at 30-days post-psilocybin session, participants reported significant reductions in depression and anxiety symptoms and significant improvements in well-being. These benefits were observed in a community-based population that included individuals with multiple psychiatric diagnoses and those actively using psychiatric medications, populations likely to be consistently included in state-regulated psilocybin frameworks. Our findings suggest that psilocybin services delivered within a structured and supportive framework can potentially produce therapeutic benefits. This translational evidence is needed to inform policy and community implementation as jurisdictions worldwide consider regulated psilocybin programs.

As a mental health intervention, psilocybin is particularly effective when combined with therapeutic support (Carhart-Harris et al., 2016, 2018; Dorczok et al., 2025; Griffiths et al., 2016), though therapeutic support is not uniformly included in studies of psilocybin’s impact on mental health (Dorczok et al., 2025). The results of this study were achieved under an enhanced, supportive model, relative to the minimum requirements set forth under Oregon’s Measure 109, through incorporating comprehensive screening, multiple preparation sessions, and structured integration support with licensed facilitators of which many held additional clinical health licenses. Such enhancements potentially influenced the overall significant improvements in mental health outcomes among study participants, including limited persistent visual perceptual or psychological aftereffects and few reports of psychedelic harm, which is consistent with prior studies (Evans et al., 2025). Research also indicates that integrative psychological support and facilitated processing after psychedelic-assisted sessions increase the likelihood of sustained benefits (Gorman et al., 2021; Greń et al., 2024).

Study participants demonstrated significant improvements in self-reported symptoms of depression, anxiety, and well-being. At 30-days post-session, 65% of participants reported normal well-being despite approximately one-third continuing to report mild to severe symptoms of depression or anxiety. Because the WHO-5 captures mental and emotional domains related to psychological flexibility and adaptive functioning (Topp et al., 2015), these findings may suggest a growing capacity for change and emerging emotional resilience, which are domains of functioning that individuals may particularly value (Connell et al., 2014). Other studies of the impacts of psilocybin on well-being likewise reported increased well-being or life satisfaction post-psilocybin session (Mans et al., 2021; Wiepking et al., 2023).

Consistent with our results, a recent naturalistic study of U.S. veterans attending psychedelic retreats outside the U.S. documented clinically significant reductions in depression with the PHQ-9 one-month post-psychedelic session, though not all participants consumed psilocybin as the psychedelic substance (some received ayahuasca) (Calnan et al., 2025). A naturalistic study from Germany similarly reported significant improvements in depression (PHQ-9) and anxiety (GAD-7) symptoms following a psilocybin session, with an approximate 25 mg psilocybin dose from fresh mushrooms, though follow-up timepoints were at 1 week and 3 months (Irrmischer et al., 2025). This study also included participants with multiple psychiatric diagnosis, which is less commonly studied (Aaronson et al., 2024; Rosenblat et al., 2024). In our study, nearly two-thirds of participants had mental health diagnoses, many concomitant, and few experienced post-session persistent aftereffects, adding to the extant evidence that psilocybin services may be safely delivered to populations experiencing a range of mental health conditions in real-world settings.

In clinical trials, dose is standardized and often fixed at 25 mg of synthetic psilocybin (Back et al., 2024; Carhart-Harris et al., 2021; Goodwin, Croal, et al., 2023; Goodwin et al., 2022; Rosenblat et al., 2024), including the recent COMPASS Pathways Phase 3 COMP005 trial (Compass, 2025). The average doses administered in our study (27.8 mg TPE, 24.3 mg psilocybin) were comparable to trials’ standard dose, and outcomes were similar whether assessed by TPE or psilocybin content alone. The novel dosing protocol described in our study, which accounts for both psilocybin and psilocin content, provides a useful framework for other states developing regulated programs utilizing natural-derived mushroom products, and may inform policy decisions regarding maximum dose limits. While Oregon’s legal maximum is 50 mg psilocybin, this study demonstrated meaningful improvements in outcomes with limited persistent aftereffects at an average dose below 30 mg TPE, suggesting that lower doses may be well-tolerated and positively improve mental health in real-world settings. However, controlled research is needed to confirm both safety and efficacy of this dosing practice, and among a greater diversity of populations than was included in this study.

Individuals taking psychiatric medications are often excluded from clinical trials to reduce the risk of a psilocybin blunting effect, allow for uniform dosing, and minimize the risk of serotonin toxicity (MacCallum et al., 2022; Malcolm and Thomas, 2022). Two psilocybin clinical trials have examined the impacts on participants taking psychiatric medications, and found reduced depressive symptoms (Goodwin, Croal, et al., 2023), and reduced adverse effects of psilocybin (Becker et al., 2022). In our observational study, we found that participants using psychiatric medications consumed modestly higher average psilocybin doses, potentially reflecting pre-emptive adjustment for anticipated psychiatric medication-related blunting (MacCallum et al., 2022). Sensitivity analyses restricted to this subgroup yielded results consistent with the main findings, suggesting concurrent psychiatric medication use did not substantially alter psilocybin response. While this may suggest that psilocybin services using whole-fruit mushroom products may be effective at improving mental health outcomes without requiring psychiatric medication discontinuation, additional controlled research examining psychiatric medications and doses, and explicitly testing moderation effects, is needed (Erritzoe et al., 2024). Additionally, as this study did not systematically collect psychiatric medication class and dose, future studies that include these data may be able to better evaluate their relationship with psilocybin.

#### Strengths and limitations

This study has several notable strengths. It represents the first systematic evaluation of mental health outcomes following psilocybin administration within the first regulated framework in the US, and the first to quantify both psilocybin and psilocin content in administered doses. This real-world study design provides initial evidence of the potential effectiveness of psilocybin in naturalistic settings among individuals with mental health complexities, thereby offering a realistic assessment of potential treatment benefits that complement traditional efficacy data (McInnes et al., 2024). Rigorous data collection and participant follow-up yielded near-complete retention, with data from only three participants excluded. Within-person pre–post assessments reduced between-person confounding, and the use of widely implemented validated instruments for depression, anxiety, and well-being supports comparison with prior studies.

Limitations of the study include its small sample size and the predominance of participants residing in Oregon, impacting its generalizability; nearly half of participants traveled from other U.S. states to access services. Participants were also disproportionately higher-income, which may reflect the cost of psilocybin services and associated travel. The study site’s scholarship program, which provided financial assistance to 58% of participants, may have partly offset this by broadening access across income levels. Nonetheless, travel, lodging, and time away from work remain potential barriers, and findings may not fully generalize to individuals with fewer financial or logistical resources. Although the simulation-based sensitivity analysis indicated adequate power to detect the primary pre-to-post changes, the relatively small sample may have limited power to detect smaller associations involving participant characteristics or dose. As such, nonsignificant covariate and dosing findings should be interpreted cautiously. However, the study was adequately powered to detect medium to large effect sizes in change in mental health symptoms, supporting the robustness of the observed results. As an observational study without a control group, causality cannot be inferred, and placebo effects, self-selection bias (such as scholarship recipient participants, though a sensitivity analysis that tested the potential influence of a scholarship on outcomes did not change the overall pattern of our findings), expectancy bias, and unmeasured confounding factors (such as influential participant lifestyle or situational changes during the study period) cannot be ruled out (Lewis et al., 2026).

Additionally, across all three mental health measures, pre-session scores reflected a community sample with generally modest mental health symptoms rather than severe clinical impairment, which may have limited the potential for improvement and influenced the observed effect sizes. Lastly, the follow-up period for the primary outcomes was limited to 30-days post-session; although this duration aligns with many clinical trials and observational studies (Calnan et al., 2025; Davis et al., 2021; Griffiths et al., 2016; Rosenblat et al., 2024), longer-term effects remain to be determined and should be investigated in future studies conducted within Oregon’s regulated model (Korthuis et al., 2024). As this study’s measurement of adverse events was limited to visual perceptual and psychological aftereffects and publicly-available adverse event reports, future studies should also include detailed reporting of adverse behavioral and medical reactions (during both the psilocybin session and follow-up period). Documenting and understanding adverse consequences of psilocybin services is critical to expanding access to this novel mental health treatment (Evans et al., 2025; Korthuis et al., 2024; Schlag et al., 2022).

### Conclusion

In this prospective, naturalistic study, we found that carefully screened participants who underwent a supervised psilocybin session within Oregon’s regulated framework and consumed naturally-derived mushroom products experienced significant improvements in depression, anxiety, and overall well-being with few persisting visual perceptual or psychological aftereffects, even among those with concurrent use of psychiatric medications and complex mental health histories. These findings provide initial evidence of the potential to deliver psilocybin safely and effectively in real-world settings, broadening the potential accessibility of psilocybin services as a tool to bridge clinical and community care. For Oregon, these data strengthen confidence in the state’s pioneering regulatory framework, offering data-driven guidance for establishing standardized practices in supportive psilocybin service frameworks, while advancing policy goals to safely expand this emerging mental health field.

### Statement of Interdisciplinarity

This study establishes a foundation for future interdisciplinary inquiry into the safety and utility of regulated psilocybin services. Future collaborations that bridge the expertise of policymakers, clinicians, and both the psychedelic and general communities will be important to ensure that emerging state frameworks are grounded in both safety and community-led practices.

## Data Availability

All data produced in the present study are available upon reasonable request to the authors

## Consent to Participate

Informed written consent was obtained from all individuals included in the study.

## Ethical Considerations

All procedures were carried out following approval from the WCG Institutional Review Board who deemed the study to be human subject’s research (approval confirmation: 45583966, March 15, 2024). All study procedures were conducted in accordance with relevant guidelines and regulations.

## Acknowledgements

We thank the participants for their time, trust, and willingness to share their experiences. We also acknowledge the valuable contributions of the Bendable Therapy team members who supported data collection: Brooke Harralson, River Jenkins, Sam Georgi, and Matt Hailey. These individuals did not receive additional compensation for these contributions.

## Notes

### Competing Interest Statement

All authors have completed the ICMJE uniform disclosure form at www.icmje.org/coi_disclosure.pdf and declare: A.G reported receipt of honoraria for presentations at University of Louisiana, Monroe. R.R. reported ownership of Aboveground Services and receipt of honoraria for consulting with Healing Advocacy Fund and for presentations with Soundmind. J.J.Q. reported owning shares in Osmind. C.M reported employment with Aboveground Services. R.C.H. reported advisor roles for Tryp Therapeutics, Osmind, Red Light Holland, and Otsuka, and receipt of honoraria for lectures, presentations, or educational events for HMP Global, Aspen Institute, Psychedelic Science IRAS, Heartmind, Clearmind, and Psychedelic Coaching Institute; no other relationships or activities could appear to have influenced the submitted work.

### Author Declarations

This study was approved as human subjects research by the Institutional Review Board of WCG (Western-Institutional Review Board-Copernicus Group).

### Summary of Updates

Updated abstract and manuscript text to reflect changes in reporting of adverse events, now termed 'aftereffects'. Added to the limitations section.

### Funding

- Jurvetson Foundation

## References

Aaronson ST, van der Vaart A, Miller T, et al. (2024) Single-Dose Synthetic Psilocybin With Psychotherapy for Treatment-Resistant Bipolar Type II Major Depressive Episodes: A Nonrandomized Open-Label Trial. JAMA Psychiatry 81(6): 555–562.

Angyus M, Osborn S, Haijen E, et al. (2024) Validation of the imperial psychedelic predictor scale. Psychological Medicine 54(12): 3539–3547.

Back AL, Freeman-Young TK, Morgan L, et al. (2024) Psilocybin Therapy for Clinicians With Symptoms of Depression From Frontline Care During the COVID-19 Pandemic: A Randomized Clinical Trial. JAMA Network Open 7(12): e2449026.

Becker AM, Holze F, Grandinetti T, et al. (2022) Acute Effects of Psilocybin After Escitalopram or Placebo Pretreatment in a Randomized, Double-Blind, Placebo-Controlled, Crossover Study in Healthy Subjects. Clinical Pharmacology and Therapeutics 111(4): 886–895.

Calnan M, Blest-Hopley G, Busch C, et al. (2025) Exploring the Therapeutic Effects of Psychedelics Administered to Military Veterans in Naturalistic Retreat Settings. Brain and Behavior 15(7): e70660.

Carhart-Harris R, Giribaldi B, Watts R, et al. (2021) Trial of Psilocybin versus Escitalopram for Depression. New England Journal of Medicine 384(15). Massachusetts Medical Society: 1402–1411.

Carhart-Harris RL, Bolstridge M, Rucker J, et al. (2016) Psilocybin with psychological support for treatment-resistant depression: an open-label feasibility study. The Lancet Psychiatry 3(7): 619–627.

Carhart-Harris RL, Bolstridge M, Day CMJ, et al. (2018) Psilocybin with psychological support for treatment-resistant depression: six-month follow-up. Psychopharmacology 235(2): 399–408.

Cohen J (1992) Quantitative Methods in Psychology: A power primer. Psychological Bulletin 112(1): 155–159.

Compass (2025) Compass Pathways Successfully Achieves Primary Endpoint in First Phase 3 Trial Evaluating COMP360 Psilocybin for Treatment-Resistant Depression. Available at: https://ir.compasspathways.com/News--Events-/news/news-details/2025/Compass-Pathways-Successfully-Achieves-Primary-Endpoint-in-First-Phase-3-Trial-Evaluating-COMP360-Psilocybin-for-Treatment-Resistant-Depression/default.aspx (accessed 12 October 2025).

Connell J, O’Cathain A and Brazier J (2014) Measuring quality of life in mental health: Are we asking the right questions? Social Science & Medicine 120: 12–20.

Davis AK, Barrett FS, May DG, et al. (2021) Effects of Psilocybin-Assisted Therapy on Major Depressive Disorder: A Randomized Clinical Trial. JAMA Psychiatry 78(5): 481–489.

de la Salle S, Kettner H, Thibault Lévesque J, et al. (2024) Longitudinal experiences of Canadians receiving compassionate access to psilocybin-assisted psychotherapy. Scientific Reports 14(1). Nature Publishing Group: 16524.

Dorczok MC, Mittmann G, Ettl T, et al. (2025) Psilocybin-assisted psychotherapy in adults with depression – A literature review. Progress in Neuro-Psychopharmacology and Biological Psychiatry 142: 111508.

Erritzoe D, Barba T, Spriggs MJ, et al. (2024) Effects of discontinuation of serotonergic antidepressants prior to psilocybin therapy versus escitalopram for major depression. Journal of Psychopharmacology (Oxford, England) 38(5): 458–470.

Evans J, Aixalà M, Anderson BT, et al. (2025) On Minimizing Risk and Harm in the Use of Psychedelics. Psychiatric Research and Clinical Practice 7(1): 4–8.

Goodwin GM, Aaronson ST, Alvarez O, et al. (2022) Single-Dose Psilocybin for a Treatment-Resistant Episode of Major Depression. New England Journal of Medicine 387(18). Massachusetts Medical Society: 1637–1648.

Goodwin GM, Croal M, Feifel D, et al. (2023) Psilocybin for treatment resistant depression in patients taking a concomitant SSRI medication. Neuropsychopharmacology 48(10): 1492–1499.

Goodwin GM, Aaronson ST, Alvarez O, et al. (2023) Single-dose psilocybin for a treatment-resistant episode of major depression: Impact on patient-reported depression severity, anxiety, function, and quality of life. Journal of Affective Disorders 327: 120–127.

Gorman I, Nielson EM, Molinar A, et al. (2021) Psychedelic Harm Reduction and Integration: A Transtheoretical Model for Clinical Practice. Frontiers in Psychology 12. Frontiers.

Greń J, Gorman I, Ruban A, et al. (2024) Call for evidence-based psychedelic integration. Experimental and Clinical Psychopharmacology 32(2): 129–135.

Griffiths RR, Johnson MW, Carducci MA, et al. (2016) Psilocybin produces substantial and sustained decreases in depression and anxiety in patients with life-threatening cancer: A randomized double-blind trial. Journal of Psychopharmacology 30(12). SAGE Publications Ltd STM: 1181–1197.

Irrmischer M, Puxty D, Yıldırım BO, et al. (2025) Moderating factors in psilocybin-assisted treatment affecting mood and personality: A naturalistic, open-label investigation. Psychopharmacology 242(4): 725–740.

Jensen ME, Stenbæk DS, Messell CD, et al. (2025) Single-dose psilocybin therapy for alcohol use disorder: Pharmacokinetics, feasibility, safety and efficacy in an open-label study. Journal of Psychopharmacology 39(5). SAGE Publications Ltd STM: 463–473.

Khan FA, Pandupuspitasari NS, Tencomnao T, et al. (2026) Breaking the chains of depression: A systematic review and meta-analysis of psilocybin therapy. Journal of Affective Disorders 397: 120882.

Korthuis PT, Hoffman K, Wilson-Poe AR, et al. (2024) Developing the Open Psychedelic Evaluation Nexus consensus measures for assessment of supervised psilocybin services: An e-Delphi study. Journal of Psychopharmacology 38(8). SAGE Publications Ltd STM: 761–768.

Kroenke K, Spitzer RL and Williams JBW (2001) The PHQ-9. Journal of General Internal Medicine 16(9): 606–613.

Lewis BR, Reid MJ, Novick AM, et al. (2026) Challenges with clinical trial participants in studies with classical psychedelics: A position statement from the National Network of Depression Centers’ task group on psychedelics and related compounds. Journal of Psychopharmacology. SAGE Publications Ltd STM: 02698811251413490.

MacCallum CA, Lo LA, Pistawka CA, et al. (2022) Therapeutic use of psilocybin: Practical considerations for dosing and administration. Frontiers in Psychiatry 13: 1040217.

Malcolm B and Thomas K (2022) Serotonin toxicity of serotonergic psychedelics. Psychopharmacology 239(6): 1881–1891.

Mans K, Kettner H, Erritzoe D, et al. (2021) Sustained, Multifaceted Improvements in Mental Well-Being Following Psychedelic Experiences in a Prospective Opportunity Sample. Frontiers in Psychiatry 12. Frontiers.

McInnes LA, Marton TF and Qian JJ (2024) Embracing pragmatism for ketamine insurance coverage: Leveraging real-world evidence. Journal of Affective Disorders 352: 199–200.

Morton E, Sakai K, Ashtari A, et al. (2023) Risks and benefits of psilocybin use in people with bipolar disorder: An international web-based survey on experiences of ‘magic mushroom’ consumption. Journal of Psychopharmacology 37(1). SAGE Publications Ltd STM: 49–60.

Nahlawi A, Ptaszek LM and Ruskin JN (2025) Cardiovascular effects and safety of classic psychedelics. Nature Cardiovascular Research 4(2): 131–144.

Nygart VA, Pommerencke LM, Haijen E, et al. (2022) Antidepressant effects of a psychedelic experience in a large prospective naturalistic sample. Journal of Psychopharmacology (Oxford, England) 36(8): 932–942.

Oregon Health Authority (2024) Public Health Division-Chapter 333, Division 333 Psilocybin, Client Information Form. Available at: https://secure.sos.state.or.us/oard/viewSingleRule.action?ruleVrsnRsn=319978 (accessed 11 October 2025).

Oregon Health Authority (2025) Public Health Division - Chapter 333. Available at: https://secure.sos.state.or.us/oard/viewSingleRule.action?ruleVrsnRsn=319968 (accessed 3 August 2026).

Oregon Health Authority (n.d.) Oregon Psilocybin Services. Available at: https://www.oregon.gov/oha/PH/PREVENTIONWELLNESS/Pages/Oregon-Psilocybin-Services.aspx (accessed 11 October 2025).

Oregon Legislature (2025) ORS 475A.372 – Requirement of psilocybin service center operator to collect, maintain, aggregate and submit to Oregon Health Authority specified information; client opt-out; exemption from disclosure; rules. Available at: https://oregon.public.law/statutes/ors_475a.372 (accessed 3 August 2026).

Oregon Psilocybin Services (2025) Oregon Psilocybin Services: Health and Safety Fact Sheet. Oregon Health Authority. Available at: https://www.oregon.gov/oha/PH/PREVENTIONWELLNESS/Documents/OPS-Health-and-Safety.pdf.

Oregon Psilocybin Services (n.d.) Oregon Psilocybin Services (OPS) Psilocybin Product Potency Information. Oregon Health Authority. Available at: https://www.oregon.gov/oha/PH/PREVENTIONWELLNESS/Documents/OPS-Psilocybin-Product-Potency-Information.pdf.

Passie T, Seifert J, Schneider U, et al. (2002) The pharmacology of psilocybin. Addiction Biology 7(4): 357–364.

Raison CL, Sanacora G, Woolley J, et al. (2023) Single-Dose Psilocybin Treatment for Major Depressive Disorder: A Randomized Clinical Trial. JAMA 330(9): 843–853.

Rosenblat JD, Meshkat S, Doyle Z, et al. (2024) Psilocybin-assisted psychotherapy for treatment resistant depression: A randomized clinical trial evaluating repeated doses of psilocybin. Med 5(3): 190–200.e5.

Ross S, Bossis A, Guss J, et al. (2016) Rapid and sustained symptom reduction following psilocybin treatment for anxiety and depression in patients with life-threatening cancer: a randomized controlled trial. Journal of Psychopharmacology 30(12). SAGE Publications Ltd STM: 1165–1180.

Schlag AK, Aday J, Salam I, et al. (2022) Adverse effects of psychedelics: From anecdotes and misinformation to systematic science. Journal of Psychopharmacology 36(3). SAGE Publications Ltd STM: 258–272.

Shin C, Lee S-H, Han K-M, et al. (2019) Comparison of the Usefulness of the PHQ-8 and PHQ-9 for Screening for Major Depressive Disorder: Analysis of Psychiatric Outpatient Data. Psychiatry Investigation 16(4): 300–305.

Spitzer RL, Kroenke K, Williams JBW, et al. (2006) A brief measure for assessing generalized anxiety disorder: the GAD-7. Archives of Internal Medicine 166(10): 1092–1097.

Tap SC, Thomas K, Páleníček T, et al. (2025) Concomitant use of antidepressants and classic psychedelics: A scoping review. Journal of Psychopharmacology. SAGE Publications Ltd STM: 02698811251368360.

Topp CW, Østergaard SD, Søndergaard S, et al. (2015) The WHO-5 Well-Being Index: a systematic review of the literature. Psychotherapy and Psychosomatics 84(3): 167–176.

von Rotz R, Schindowski EM, Jungwirth J, et al. (2023) Single-dose psilocybin-assisted therapy in major depressive disorder: a placebo-controlled, double-blind, randomised clinical trial. eClinicalMedicine 56: 101809.

Wiepking L, Bruin E de and Ghiţă A (2023) The potential of psilocybin use to enhance well-being in healthy individuals – A scoping review. Journal of Psychedelic Studies 7(3): 184–199.

Zhou K, de Wied D, Carhart-Harris RL, et al. (2025) Prediction of hallucinogen persisting perception disorder and thought disturbance symptoms following psychedelic use. PNAS Nexus 4(4): pgae560.

