## Appendix A for "Prospective real-world evidence of mental health outcomes following supervised psilocybin services within Oregon’s state-regulated model"

**Contents:**

1. Supplemental Table 1: Summary of validated questionnaires used in the study
2. Supplemental Table 2: Changes in PHQ-8, GAD-7, and WHO-5 scores from pre-session to 1-day post-session (n=88 participants)
3. Supplemental Table 3: Changes in PHQ-8, GAD-7, and WHO-5 scores from pre-session to 30-days post-session controlling for dose measured in psilocybin (n=88 participants)
4. Supplemental Table 4: Changes in PHQ-8, GAD-7, and WHO-5 scores from pre-session to 30-days post-session among participants who completed a single administration session (n=75 participants)
5. Supplemental Table 5: Changes in PHQ-8, GAD-7, and WHO-5 scores from pre-session to 30-days post-session comparing those with concurrent psychiatric medication use (n=41 participants) with those without (n=47 participants)
6. Supplemental Table 6: Item-level frequencies of visual perceptual aftereffects reported in post-session surveys
7. Supplemental Table 7: Univariate regression models examining predictors of session dose measured in TPE (mg, n=88 participants)
8. Supplemental Table 8: Associations between dose (TPE) and predictors from the Imperial Psychedelic Predictor Scale (iPPS, n=88 participants)

**Supplemental Table 1:** Summary of validated questionnaires used in the study^a^

| **Scale** | **Name** | **Topic** | **Scoring** | **Scale** | **Diagnostic Threshold** | **Directionality** | **Reliability within study sample^b^** |
| --- | --- | --- | --- | --- | --- | --- | --- |
| PHQ-8^c^ | Patient Health Questionnaire-8 | Depression | 0-24 | 24 | 10 | Higher score indicates greater symptom severity | ɑ=0.87 all time points |
| GAD-7 | Generalized Anxiety Disorder-7 questionnaire | Anxiety | 0-21 | 21 | 10 | Higher score indicates greater symptom severity | ɑ=0.91 (pre-session); ɑ=0.94 (1-day post-session); ɑ=0.89 (30-days post-session) |
| WHO-5 | World Health Organization Well-Being Index | Well-Being | 0-25 | 25 | 13 | Lower scores indicate poor well-being | ɑ=0.90 (pre-session); ɑ=0.89 (1-day post-session); ɑ=0.93 (30-days post-session) |
| iPPS | Imperial  Psychedelic Predictor Scale | Psychedelic Prepared-  ness | 0-100 | 100 | N/A | Higher scores indicate greater preparedness | N/A |
| HPPD^d^ | Hallucinogen Persisting Perception Disorder | Hallucinogen Persisting Perception Disorder | Yes/No | N/A | Responding “Yes” to Item 2 and “Not True” to Item 3 may indicate HPPD | Responding “Yes” to Item 2 and “Not True” to Item 3 may indicate HPPD | N/A |

^a^ Abbreviations: GAD-7, Generalized Anxiety Disorder-7 questionnaire; HPPD, Hallucinogen Persisting Perception Disorder; iPPS, Imperial Psychedelic Predictor Scale; PHQ-8, Patient Health Questionnaire-8; WHO-5, World Health Organization Well-Being Index

^b^ Measured as Cronbach’s alpha.

^c^ This study modified the standard PHQ-9 to the PHQ-8 by excluding the suicidal ideation item; scores were adjusted from a total of 27 (PHQ-9) to 24 (PHQ-8).

^d^ HPPD *Item 1* asks if one or more of the following symptoms are being re-experienced since the psychedelic session (participants selected all that applied): geometric hallucinations, false perceptions of movement in peripheral visual fields, flashes of colors, intensified colors, trails of images of moving objects, positive after-images, halos around objects, macropsia (objects appear larger than they normally are), micropsia (objects appear smaller than they normally are), none of the above. If any symptoms were selected in Item 1, *Item 2* asks (yes or no): do these cause significant distress or impairment in social, occupational, or other important areas of functioning? If Item 2 is yes, *Item 3* asks (yes or no, updated to True/Not True in the study): These symptoms are not due to a general medical condition (e.g., anatomical lesions and infections of the brain, visual epilepsies) and are not better accounted for by another mental disorder (e.g., delirium, dementia, Schizophrenia) or hypnopompic hallucinations.

**Supplemental Table 2:** Changes in PHQ-8, GAD-7, and WHO-5 scores from pre-session to 1-day post-session (n=88 participants)^a^

| **Measure** | **Pre-Session M (SE)** | **1-day post session M (SE)** | **Change pre-session to post-session**  **M (SE) [95% CI]** | **p-value** | **t (df)** | **Cohen’s d [95% CI]** |
| --- | --- | --- | --- | --- | --- | --- |
| PHQ-8 | 9.15 (0.60) | 7.14 (0.60) | -2.01 (0.70) [-3.41, -0.61] | 0.005 | -2.86 (87) | -0.36 [-0.61, -0.11] |
| GAD-7 | 8.22 (0.61) | 7.10 (0.61) | -1.11 (0.65) [-2.40, 0.17] | 0.09 | -1.72 (87) | -0.19 [-0.42, 0.03] |
| WHO-5 | 11.2 (0.54) | 12.9 (0.54) | +1.70 (0.60) [0.52, 2.89] | 0.005 | 2.86 (87) | 0.34 [0.10, 0.58] |

^a^ Results reported as mean (SE). Covariates included birth sex (male/female/decline to answer), age (years), concurrent psychiatric medication use (yes/no), and TPE (mg) as a continuous dosage. In the PHQ-8 and WHO-5 models, the “Decline to Answer” category differed significantly from the Female reference group (p=0.02 and p=0.04, respectively); however, these estimates were based on a single participant and should not be interpreted as evidence of a birth-sex effect. TPE was a significant covariate in the WHO-5 model (p=0.04). No covariates were significant in the GAD-7 model, and no other covariates were significant in the PHQ-8 or WHO-5 models. Cohen’s d interpretations: small d=0.20-0.49; moderate d=0.50-0.79; large d≥0.80. Abbreviations: GAD-7, Generalized Anxiety Disorder-7 questionnaire; PHQ-8, Patient Health Questionnaire-8; WHO-5, World Health Organization Well-Being Index.

**Supplemental Table 3:** Changes in PHQ-8, GAD-7, and WHO-5 scores from pre-session to 30-days post-session controlling for dose measured in psilocybin (n=88 participants)^a^

| **Measure** | **Pre-Session M (SE)** | **30-days post session (30-days) M (SE)** | **Change pre-session to post-session**  **M (SE) [95% CI]** | **p-value** | **t(df)** | **Cohen’s d [95% CI]** |
| --- | --- | --- | --- | --- | --- | --- |
| PHQ-8 | 9.15 (0.55) | 4.52 (0.55) | -4.63 (0.57) [-5.77, -3.48] | <0.001 | -8.06 (87) | -0.90 [-1.12, -0.68] |
| GAD-7 | 8.22 (0.50) | 3.36 (0.50) | -4.85 (0.57) [-5.99, -3.71] | <0.001 | -8.48 (87) | -1.04 [-1.29, -0.80] |
| WHO-5 | 11.2 (0.54) | 15.5 (0.54) | +4.26 (0.55) [3.18, 5.35] | <0.001 | 7.80 (87) | 0.84 [0.62, 1.05] |

^a^ Results reported as mean (SE). Covariates included birth sex (male/female/decline to answer), age (years), concurrent psychiatric medication use (yes/no), and psilocybin (mg) as a continuous dosage. For the model with PHQ-8, concurrent use of mental health medications (p=0.038) was positively associated with PHQ-8 scores. For the model with GAD-7, age was negatively associated with 30-day post-session GAD-7 scores (p=0.048) and concurrent psychiatric medication use was positively associated with 30-day post-session GAD-7 scores (p= 0.047). No covariates were significant in the model with WHO-5. Cohen’s d interpretations: small d=0.20-0.49; moderate d=0.50-0.79; large d≥0.80. Abbreviations: GAD-7, Generalized Anxiety Disorder-7 questionnaire; PHQ-8, Patient Health Questionnaire-8; WHO-5, World Health Organization Well-Being Index.

**Supplemental Table 4:** Changes in PHQ-8, GAD-7, and WHO-5 scores from pre-session to 30-days post-session among participants who completed a single administration session (n=75 participants)^a^

| **Measure** | **Pre-session M (SE)** | **30-days post session M (SE)** | **Change pre-session to post-session**  **M (SE) [95% CI]** | **p-value** | **t(df)** | **Cohen’s d [95% CI]** |
| --- | --- | --- | --- | --- | --- | --- |
| PHQ-8 | 9.11 (0.60) | 4.57 (0.60) | -4.53 (0.62) [-5.77, -3.30] | <0.001 | -7.32 (74) | -0.87 [-1.10, -0.63] |
| GAD-7 | 8.07 (0.54) | 3.33 (0.54) | -4.73 (0.61) [-5.96, -3.51] | <0.001 | -7.72 (74) | -1.01 [-1.27, -0.75] |
| WHO-5 | 10.9 (0.59) | 15.3 (0.59) | +4.37 (0.61) [3.16, 5.59] | <0.001 | 7.17 (74) | 0.86 [0.62, 1.09] |

^a^ Results reported as mean (SE). Covariates included birth sex (male/female), age (years), concurrent psychiatric medication use (yes/no), and TPE (mg) as a continuous dosage. No covariates were significant in the PHQ-8 or WHO-5 models; concurrent psychiatric medication use was positively associated with GAD-7 scores (p=0.020). Cohen’s d interpretations: small d=0.20-0.49; moderate d=0.50-0.79; large d≥0.80. Abbreviations: GAD-7, Generalized Anxiety Disorder-7 questionnaire; PHQ-8, Patient Health Questionnaire-8; WHO-5, World Health Organization Well-Being Index.

**Supplemental Table 5:** Changes in PHQ-8, GAD-7, and WHO-5 scores from pre-session to 30-days post-session comparing those with concurrent psychiatric medication use (n=41 participants) with those without (n=47 participants)^a^

| **Measure** | **Pre-session M (SE)** | **30-days post-session M (SE)** | **Change pre-session to post-session**  **M (SE) [95% CI]** | **p-value** | **t(df)** | **Cohen’s d [95% CI]** |
| --- | --- | --- | --- | --- | --- | --- |
| Participants with concurrent psychiatric medication use | | | | | | |
| PHQ-8 | 9.90 (0.89) | 5.54 (0.89) | -4.37 (0.89) [-6.16, -2.57] | <0.001 | -4.91 (40) | -0.77 [-1.08, -0.45] |
| GAD-7 | 8.68 (0.78) | 4.44 (0.78) | -4.24 (0.81) [-5.87, -2.62] | <0.001 | -5.27 (40) | -0.86 [-1.18, -0.53] |
| WHO-5 | 10.8 (0.83) | 14.5 (0.83) | 3.71 (0.85) [1.98, 5.44] | <0.001 | 4.34 (40) | 0.70 [0.37, 1.02] |
| Participants without concurrent psychiatric medication use | | | | | | |
| PHQ-8 | 8.49 (0.69) | 3.64 (0.69) | -4.85 (0.75) [-6.36, -3.34] | <0.001 | -6.46 (46) | -1.03 [ -1.36, -0.71] |
| GAD-7 | 7.81 (0.64) | 2.43 (0.64) | -5.38 (0.81) [-7.01, -3.75] | <0.001 | -6.65 (46) | -1.22 [-1.59, -0.85] |
| WHO-5 | 11.6 (0.72) | 16.3 (0.72) | 4.74 (0.70) [3.33, 6.16] | <0.001 | 6.77 (46) | 0.97 [0.68, 1.25] |

^a^ Results reported as mean (SE). Covariates included birth sex (male/female/decline to answer), age (years), and total psilocybin equivalents (TPE, mg) as a continuous dosage. No covariates were significant in any of the models. Cohen’s d interpretations: small d=0.20-0.49; moderate d=0.50-0.79; large d≥0.80. Abbreviations: GAD-7, Generalized Anxiety Disorder-7 questionnaire; PHQ-8, Patient Health Questionnaire-8; WHO-5, World Health Organization Well-Being Index.

**Supplemental Table 6: Item-level frequencies of visual perceptual aftereffects reported in post-session surveys**

| **Perceptual symptom** | **1-day**  **N (%)** | **30-day**  **N (%)** |
| --- | --- | --- |
| None of the above | 53 (60.2) | 72 (81.8) |
| Intensified colors | 19 (21.6) | 6 (6.8) |
| False perceptions of movement in the peripheral visual fields | 14 (15.9) | 7 (8.0) |
| Flashes of colors | 10 (11.4) | 3 (3.4) |
| Geometric hallucinations | 10 (11.4) | 5 (5.7) |
| Trails of images of moving objects | 9 (10.2) | 4 (4.5) |
| Positive afterimages | 7 (8.0) | 4 (4.5) |
| Halos around objects | 6 (6.8) | 1 (1.1) |
| Macropsia (objects appear larger than they actually are) | 4 (4.5) | 0 (0) |
| Micropsia (objects appear smaller than they actually are) | 1 (1.1) | 0 (0) |

**Supplemental Table 7:** Univariate regression models examining predictors of session dose measured in TPE (mg, n=88 participants)^a^

| **Predictor** | **Estimate** | **Std Error** | **t-value** | **p-value** |
| --- | --- | --- | --- | --- |
| Pre-Session GAD-7 | 0.12 | 0.16 | 0.77 | 0.44 |
| Pre-Session PHQ-8 | -0.007 | 0.15 | -0.05 | 0.96 |
| Pre-Session WHO-5 | -0.13 | 0.17 | -0.81 | 0.42 |
| Age | 0.04 | 0.06 | 0.61 | 0.55 |
| Sex (Male) | 0.70 | 1.76 | 0.40 | 0.69 |
| Sex (Decline to Answer) | -7.52 | 8.30 | -0.91 | 0.37 |
| Complaint (Depression) | 3.09 | 1.72 | 1.80 | 0.08 |
| Complaint (Anxiety) | -0.93 | 1.77 | -0.53 | 0.6 |
| Complaint (PTSD) | -0.08 | 2.22 | -0.04 | 0.97 |
| Currently Taking Psychiatric Meds (Yes) | 3.44 | 1.71 | 2.01 | 0.05* |
| Prior Psychedelic Use | -0.17 | 1.83 | -0.10 | 0.93 |

^a^ Abbreviations: GAD-7, Generalized Anxiety Disorder-7 questionnaire; PHQ-8, Patient Health Questionnaire-8; TPE, total psilocybin equivalents; WHO-5, World Health Organization Well-Being Index.

**Supplemental Table 8:** Associations between dose (TPE) and predictors from the Imperial Psychedelic Predictor Scale (iPPS, n=88)^a^

| **Predictor** | **Estimate** | **Std Error** | **t-value** | **p-value** |
| --- | --- | --- | --- | --- |
| (Intercept) | 15.8 | 12.1 | 1.31 | 0.19 |
| Surrender | -0.11 | 0.09 | -1.18 | 0.24 |
| Open | 0.0003 | 0.17 | 0.002 | 0.999 |
| Prepared | 0.20 | 0.10 | 2.07 | 0.04* |
| Comfortable | 0.08 | 0.10 | 0.72 | 0.47 |
| Mood | -0.10 | 0.09 | -1.12 | 0.26 |
| Anxious | 0.007 | 0.03 | 0.25 | 0.81 |
| Intention | -0.07 | 0.06 | -1.05 | 0.30 |
| Relationship | 0.12 | 0.14 | 0.87 | 0.39 |
| Feeling^b^ | 0.05 | 0.18 | 0.28 | 0.78 |

^a^ iPPS subscales were entered simultaneously into the multivariable regression model to assess their independent contributions to TPE (mg). Abbreviations: iPPS, Imperial Psychedelic Predictor Scale; TPE, total psilocybin equivalents.

^b^ In the iPPS, the ‘Feeling’ variable is only included in analysis for individuals undergoing a group psilocybin session, n=38 participants in this study.
